# Pharmacokinetics/pharmacodynamics by Race: Analysis of a Peginterferon β-1a Phase I Study

**DOI:** 10.1101/2022.04.01.22272847

**Authors:** Yuan Zhao, Oksana Mokliatchouk, Nancy F Ramia, Maria L Naylor, Cherié L Butts

**Affiliations:** Biogen, Cambridge, MA, 02142, USA

**Keywords:** multiple sclerosis, peginterferon beta-1a, pharmacokinetics, pharmacodynamics, underserved populations

## Abstract

**Background:** Black/African-American participants are underrepresented in clinical trials but can experience a greater burden of disease, such as multiple sclerosis, than other racial groups in the United States. A phase 1, open-label, 2-period crossover study that demonstrated bioequivalence of subcutaneous (SC) and intramuscular (IM) injection of peginterferon beta-1a in healthy volunteers enrolled a similar proportion of Black/African-American and White participants, enabling a subgroup analysis comparing these groups.

**Methods:** Peginterferon beta-1a 125 μg was administered by SC or IM (1:1) injection, followed by a 28-day washout period before a second injection using the alternate method. Primary endpoints were maximum observed concentration (C_max_) and area under the concentration-time curve from Hour 0 to infinity (AUC_inf_). Secondary endpoints included safety, tolerability, and additional pharmacokinetic and pharmacodynamic parameters.

**Findings:** This analysis included 70 (51.5%) Black and 59 (43.3%) White participants. Black participants exhibited a 29.8% higher geometric mean C_max_ of peginterferon beta-1a than White participants following SC administration and demonstrated similar values following IM administration. Black participants displayed 31.0% versus 11.8% higher geometric mean AUC_inf_ values than White participants with SC versus IM administration. Neopterin dynamics and safety signals were similar between groups, with numerically fewer adverse events reported among Black participants.

**Conclusions:** No clinically meaningful differences were identified between Black and White participants in pharmacokinetics/pharmacodynamics or safety related to peginterferon beta-1a administration, indicating that no change in dosing regimen is warranted for Black patients with MS.

**Funding:** Funding for medical writing support was provided by Biogen Inc. (Cambridge, MA, USA).

## INTRODUCTION

Certain racial and ethnic populations in the United States (US), such as those who identify as Black, are underrepresented in clinical studies.^1-3^ This includes studies of multiple sclerosis (MS).^4, 5^ According to a recent examination of 56.6 million US electronic health records, the prevalence of MS per 100,000 was 283.7 versus 226.1 for White versus Black patients.^6^ The Black population in the US is disproportionately affected by MS, with incidence rates of ∼10–12 per 100,000, compared with ∼7–9 per 100,000 among the White population.^7, 8^ However, clinical trial populations of patients with MS predominantly comprise White participants, commonly with <15% of participants self-identifying as Black.^9-12^ Low representation of Black participants in clinical trials of treatments for MS restricts providers’ and patients’ ability to assess risks and benefits of treatments in these populations, as medications may have distinct safety and efficacy profiles across different patient groups.^13, 14^ Such disparities in clinical trial recruitment, and their consequences for treatment, have prompted calls to action from federal agencies, journal editorial boards, and physicians to increase diversity in clinical trial populations.^14-19^

Differences in severity and treatment response may exist between Black and White patients with MS. Some studies have suggested Black patients with MS in the US have more severe disease at presentation^20, 21^ and a more aggressive disease course^21-25^ compared with White patients, although further evaluation is required for confirmation. Studies of MS disease modifying therapies (DMTs) in Black patients are limited, though published reports suggest that Black patients with MS have a less favorable treatment response to certain DMTs, including beta interferons, compared with White patients.^9, 11, 26^ Unfortunately, the low proportion of Black patients included in these studies and limited details on the extended disease course make it difficult to determine how well these results reflect the broader Black population.

Within the MS patient population, it is still unclear whether there are clinically significant differences in DMT pharmacokinetics (PK) and pharmacodynamics (PD) between Black and White patient populations. To establish a dosing regimen that achieves both sufficient exposure and acceptable safety in all relevant patient populations, an understanding of the PK/PD properties of treatments in these populations is essential.

Peginterferon beta-1a, a pegylated form of interferon beta-1a,^27^ administered by subcutaneous (SC) injection of 125 μg every 2 weeks (Q2W), was approved in 2014 for treatment of patients with relapsing forms of MS. Intramuscular (IM) administration was approved in 2021^28^ based on demonstration of bioequivalence of PK and PD between SC and IM administration in healthy adult participants.^29^ As of June 30 2021, peginterferon beta-1a has been prescribed to approximately 70,487 patients globally, corresponding to 141,246 person-years of exposure. In the peginterferon beta-1a bioequivalence study, approximately half of the participants self-reported as Black/African-American (70 of 136 [51.5%]). A post hoc analysis was undertaken to provide a detailed examination of PK and PD parameters and safety outcomes in Black and White participants who received peginterferon beta-1a administered by SC or IM injection in the bioequivalence study.

## METHODS

### Study Design

Details of the phase 1, open-label, multicenter, 2-period crossover study of peginterferon beta-1a in healthy adult volunteer participants have been previously reported.^29^ Briefly, participants were randomized 1:1 to receive either a single dose of IM peginterferon beta-1a 125 μg or SC peginterferon beta-1a 125 μg in treatment period 1, followed by a single dose of SC or IM peginterferon beta-1a 125 μg in treatment period 2 with a 28-day washout period between administrations.

The study was open-label, and neither the participants nor investigators were blinded. The study protocol was approved by an independent review board at each site, and the study was conducted in accordance with the International Council for Harmonisation Guidelines on Good Clinical Practice and the World Medical Association Declaration of Helsinki. All participants provided informed consent prior to evaluation for eligibility.

### Participants

Study inclusion criteria have been previously presented.^29^ All included participants were selected in keeping with standard regional practices for early-phase clinical pharmacology studies. Participant-provided information on race was collected during screening, in accordance with US Food and Drug Administration guidance on clinical trial reporting. Participants self-identified as belonging to 1 or more of the following: American Indian or Alaska Native, Asian, Black or African American, Native Hawaiian or Other Pacific Islander, White, or Other. This post hoc analysis focused on Black and White participants only as the other groups had numbers of participants too small to include.

### Endpoints

The primary endpoint for this analysis was assessment PK parameters, including maximum observed concentration (C_max_) and area under the concentration-time curve (AUC) from Hour 0 extrapolated to infinity (AUC_inf_) in Black and White study participants following SC and IM administration of peginterferon beta-1a. Secondary endpoints included incidence of adverse events (AEs), treatment-emergent AEs (TEAEs), and serious AEs (SAEs), and the assessment of secondary PK endpoints, including AUC from Time 0 to 336 hours post dose (AUC_0–336_), AUC from Time 0 to 504 hours post dose (AUC_0–504_), time to maximum observed plasma concentration (T_max_), elimination half-life (t_½_), terminal elimination rate constant (λ_Z_), and apparent total body clearance (CL/F) in the same study subgroups. Exploratory endpoints were assessment of PD parameters based on serum levels of neopterin, a marker of type 1 interferon receptor activation,^30^ and included baseline concentration (E_baseline_), area under the effect-time curve from Time 0 to 336 and 504 hours post dose (E_AUC0–336_ and E_AUC0–504_, respectively), maximum effect (E_peak_), and time to reach maximum effect (E_Tmax_).

### Statistical Analyses

Sample size calculations were not performed for this analysis. The full analysis population was defined as all randomized Black and White participants. PK was assessed in the PK population, defined as members of the full analysis population who received both doses of peginterferon beta-1a with sufficient PK measurements to allow calculation of PK parameters. PD endpoints were assessed in the PD population, which was defined as members of the full analysis population who received at least 1 dose of peginterferon beta-1a with at least 1 PD measurement after baseline. Safety outcomes were assessed in the safety population, which was defined as members of the full analysis population who received at least 1 dose of SC or IM peginterferon beta-1a. All PK, safety, and PD outcomes were analyzed using descriptive statistics. All statistical analyses were performed using SAS version 9.4 (SAS Institute, Cary, NC, USA).

## RESULTS

### Participants

The bioequivalence study was conducted between December 2018 and May 2019 at 2 US sites (Daytona Beach, FL, and Dallas, TX) and included 136 randomized, healthy volunteer participants.^29^ For these post hoc analyses, the full analysis population included 70 (51.5%) Black and 59 (43.4%) White participants (Table 1). Baseline characteristics of the 2 subgroups were similar, although Black participants tended to be younger than White participants (mean ± standard deviation [SD]: 37.2 ± 9.05 vs 43.9 ± 9.08 years). Peginterferon beta-1a PK/PD by Race

**Table 1.** Baseline Characteristics of Included Participants by Treatment (Full Analysis Population).

| Characteristic* | Black |  |  | White |  |  |
| --- | --- | --- | --- | --- | --- | --- |
|  | Peginterferon<br>beta-1a 125 µg<br>IM/peginterferon<br>beta-1a 125 µg<br>SC<br>(n = 34) | Peginterferon<br>beta-1a 125 µg<br>SC/peginterferon<br>beta-1a 125 µg IM<br>(n = 36) | Total<br>(n = 70) | Peginterferon<br>beta-1a 125 µg<br>IM/peginterferon<br>beta-1a 125 µg<br>SC<br>(n = 29) | Peginterferon<br>beta-1a 125 µg<br>SC/peginterferon<br>beta-1a 125 µg IM<br>(n = 30) | Total<br>(n = 59) |
| Age—years | 38.1 ± 10.02 | 36.3 ± 8.07 | 37.2 ± 9.05 | 44.6 ± 9.12 | 43.3 ± 9.15 | 43.9 ± 9.08 |
| Age range—years, no. (%) |  |  |  |  |  |  |
| 18 to <25 | 3 (8.8) | 3 (8.3) | 6 (8.6) | 0 | 2 (6.7) | 2 (3.4) |
| 25 to <40 | 16 (47.1) | 19 (52.8) | 35 (50.0) | 7 (24.1) | 5 (16.7) | 12 (20.3) |
| 40 to 55 | 15 (44.1) | 14 (38.9) | 29 (41.4) | 22 (75.9) | 23 (76.7) | 45 (76.3) |
| Female, no. (%) | 15 (44.1) | 10 (27.8) | 25 (35.7) | 17 (58.6) | 16 (53.3) | 33 (55.9) |
| Ethnicity, no. (%) |  |  |  |  |  |  |
| Hispanic/Latino | 4 (11.8) | 3 (8.3) | 7 (10.0) | 21 (74.2) | 17 (56.7) | 38 (64.4) |
| Not Hispanic/Latino | 30 (88.2) | 46 (91.7) | 63 (90.0) | 8 (27.6) | 13 (43.3) | 21 (35.6) |
| Height—cm | 172.6 ± 7.38 | 176.5 ± 11.38 | 174.6 ± 9.77 | 165.4 ± 10.31 | 168.8 ± 8.55 | 167.1 ± 8.61 |
| Weight—kg | 81.3 ± 10.68 | 83.1 ± 13.39 | 82.2 ± 12.09 | 72.6 ± 10.31 | 73.4 ± 10.49 | 73.0 ± 10.32 |
| BMI—kg/m<sup>2</sup> | 27.2 ± 2.32 | 26.6 ± 2.70 | 26.9 ± 2.52 | 26.5 ± 2.72 | 25.8 ± 2.98 | 26.1 ± 2.85 |

\*Plus–minus values are means ± standard deviation.
BMI, body mass index; IM, intramuscular; SC, subcutaneous.

### Pharmacokinetics and Pharmacodynamics

Following SC administration of peginterferon beta-1a, Black participants exhibited 29.8% higher geometric mean C_max_ concentration of peginterferon beta-1a (1.35 ng/mL [coefficient of variance (CV) 69.8%] vs 1.04 ng/mL [CV 86.4%]) than White participants (Figure 1A; Table S1). After IM administration, Black and White participants displayed equivalent geometric mean C_max_ concentrations of peginterferon beta-1a (1.29 ng/mL [CV 64.7%] vs 1.29 ng/mL [CV 56.8%]) (Figure 1B; Table S1), although Black participants had a 7.8% higher median concentration than White participants (1.38 [range 0.14–5.14] ng/mL vs 1.28 [range 0.12–3.51 ng/mL] (Table S1).

**Figure 1.**
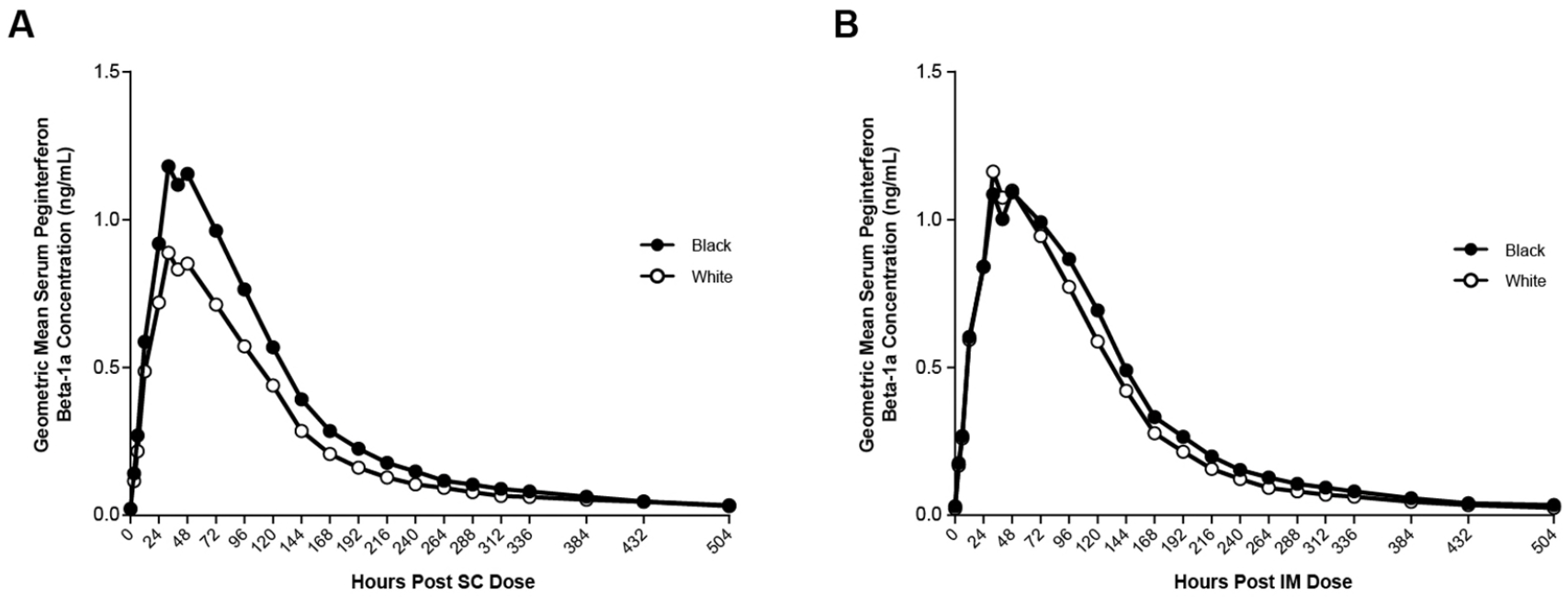
Serum Concentration Profiles for Peginterferon Beta-1a in Black and White Participants Following Single 125-μg (A) SC or (B) IM Doses (Safety Population). Baseline safety populations for SC and IM administration included 4 and 2 Black participants, respectively, and 2 White participants (IM subgroup only; baseline serum concentration missing for White SC subgroup). For all post dose time points, the safety populations for SC and IM administration ranged from 45–69 and 36–66, respectively, for Black participants, and from 31–58 and 25–59, respectively, for White participants. IM, intramuscular; SC, subcutaneous.

Black participants had 31.0% higher geometric mean AUC_inf_ with SC administration (177.24 h × ng/mL [CV 49.6%] vs 135.31 h × ng/mL [CV 52.5%]) and 11.8% higher with IM administration (176.87 h × ng/mL [CV 50.4%] vs 158.20 h × ng/mL [CV 46.6%]) compared with White participants (Table S1).

Assessments of the secondary PK endpoints, AUC_0–336_ and AUC_0–504_, paralleled the primary AUC_inf_ findings, with Black participants showing 28.8%–28.9% larger geometric mean values with SC administration (AUC_0–336_: 152.96 h × ng/mL [CV 60.35%] vs 118.71 h × ng/mL [CV 63.02%]; AUC_0–504_: 163.28 h × ng/mL [CV 58.03%] vs 126.82 h × ng/mL [CV 59.26%]) and 6.7%–8.1% larger geometric mean values with IM administration (AUC_0–336_: 160.71 h × ng/mL [CV 56.26%] vs 150.75 h × ng/mL [CV 47.69%]; AUC_0–504_: 169.03 h × ng/mL [CV 55.32%] vs 156.29 h × ng/mL [CV 46.91%]) compared with White participants (Table S1). In addition, Black participants had 8.9%–13.1% longer median peginterferon beta-1a half-life with SC administration (94.0 [range 46.1–239.9] hours vs 86.3 [range 29.1–183.5]) hours and IM administration (78.8 [range 26.3–233.3] hours vs median 69.7 [40.1–127.7] hours) compared with White participants (Table S1). Black participants had slower mean apparent total plasma clearance following SC administration (0.873 L/hour [CV 87.7%] vs 1.110 L/hour [CV 70.1%]) than White participants, although clearance was similar following IM administration (0.851 L/hour [CV 77.7%] vs 0.881 L/hour [CV 55.9%]).

Baseline and peak neopterin concentrations, time to maximum neopterin concentration, and effect AUCs were similar for Black and White study participants following SC or IM peginterferon beta-1a (Table S2). Profiles of serum neopterin concentration up to 504 hours (21 days) post dose were similar in both participant subgroups after SC or IM administration (Figure 2).

**Figure 2.**
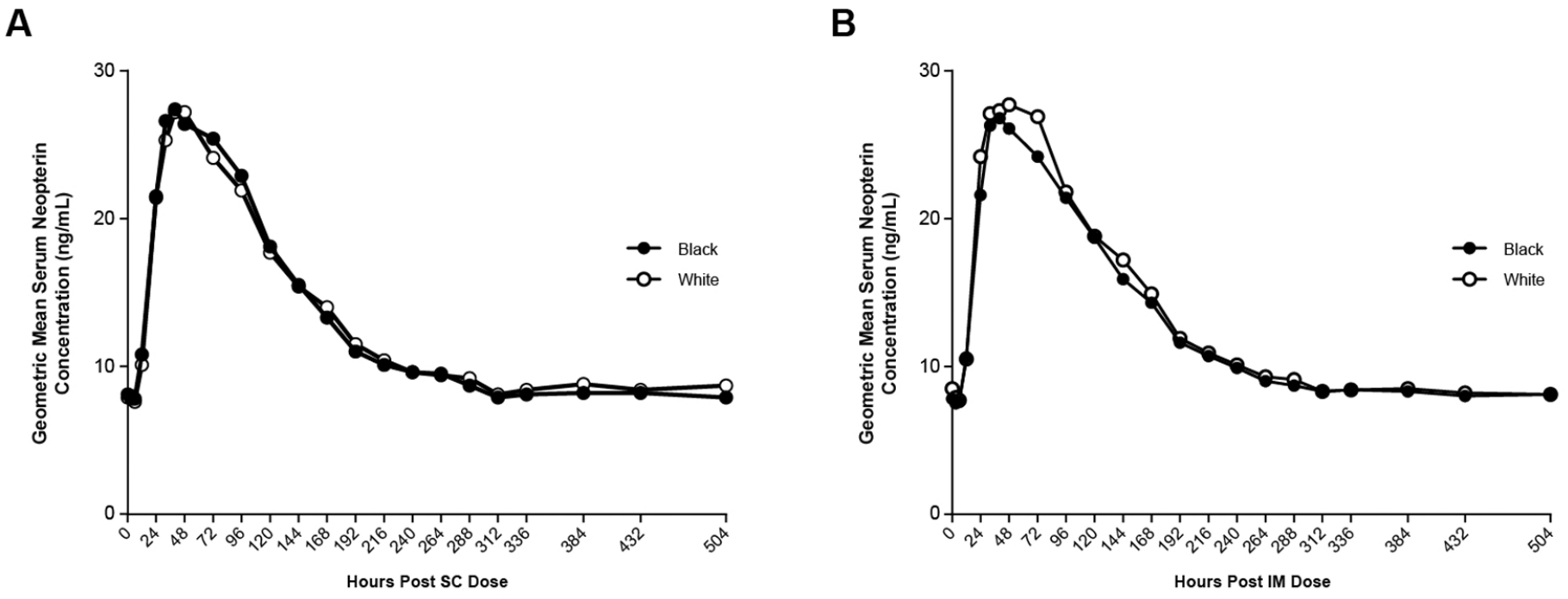
Serum Concentration Profiles for Neopterin in Black and White Participants Following Single 125-μg (A) SC and (B) IM Doses of Peginterferon Beta-1a (PD Population). For all time points, the PD populations for SC and IM administration ranged from 66–69 and 65–66, respectively, for Black participants, and 57–58 and 58–59, respectively, for White participants. IM, intramuscular; PD, pharmacodynamic; SC, subcutaneous.

### Safety

The Black safety population comprised 69 and 66 participants who received SC and IM peginterferon beta-1a, respectively, while the White safety population comprised 58 and 59 participants who received SC and IM peginterferon beta-1a, respectively (Table 2). A numerically lower incidence of TEAEs were reported by Black versus White participants who received SC (63.8% [44 of 69] vs 75.9% [44 of 58], respectively) or IM administration (54.5% [36 of 66] vs 79.7% [47 of 59], respectively) (Table 2). There were no SAEs reported by participants in either group.

**Table 2.**
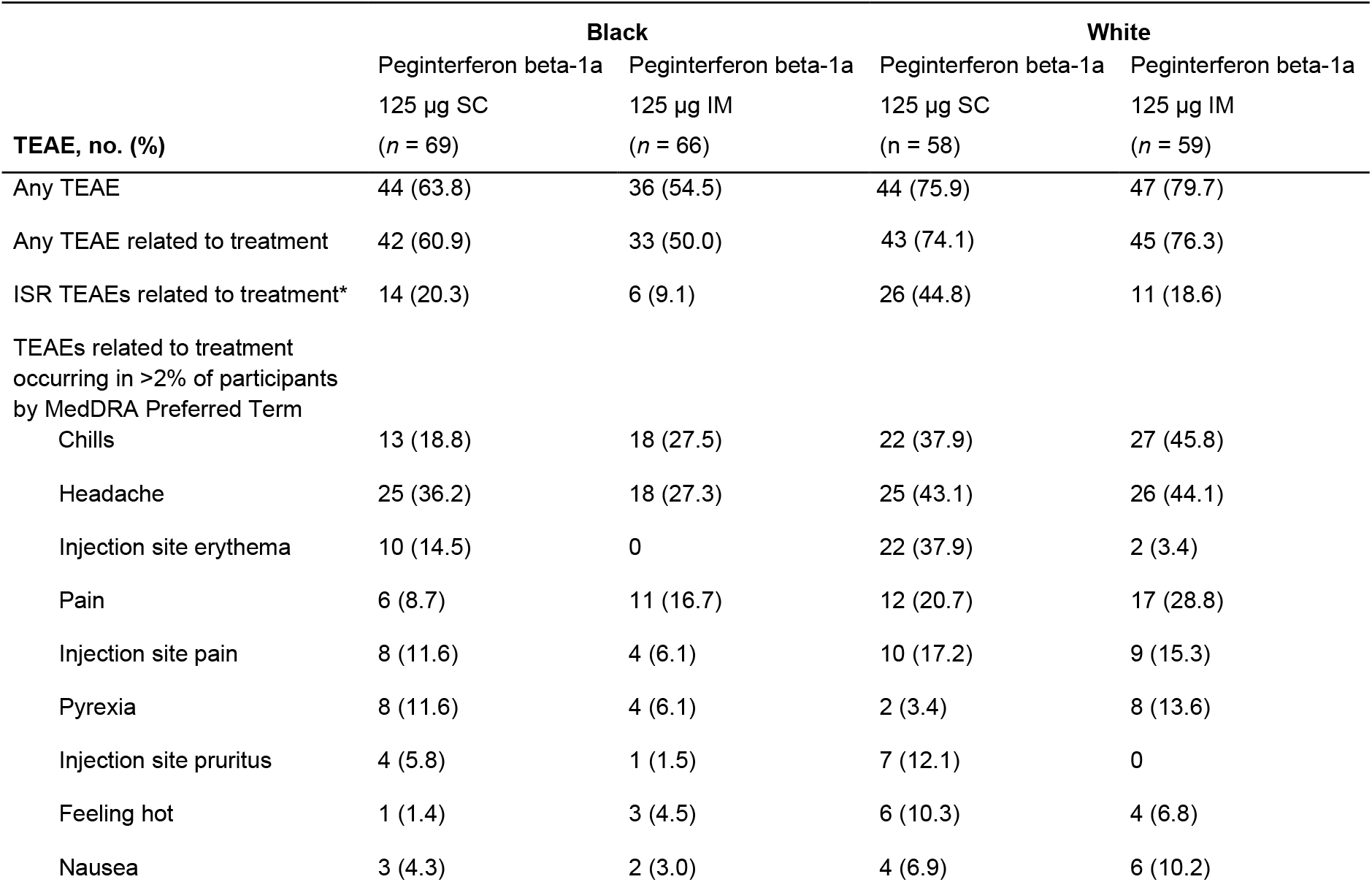
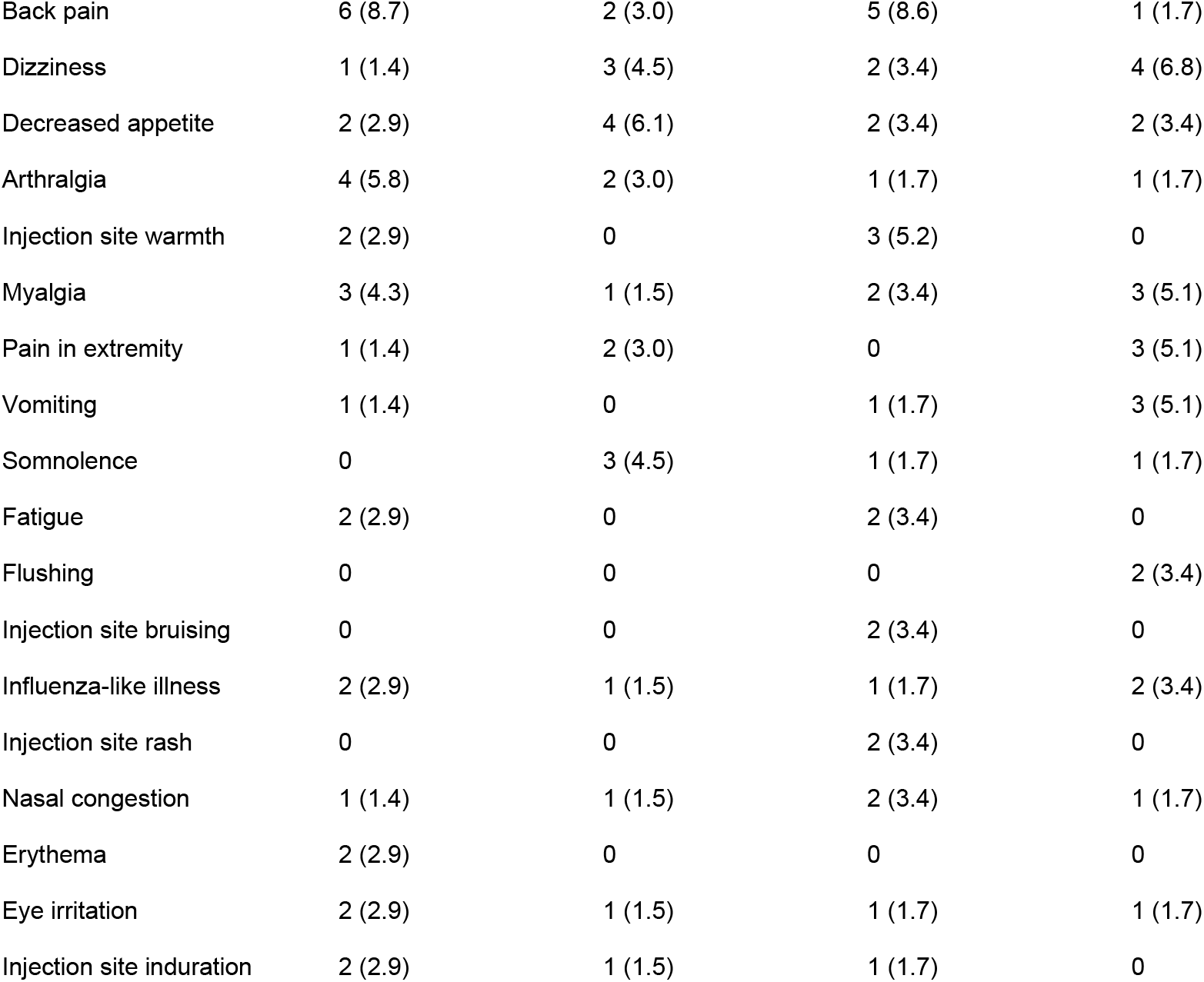
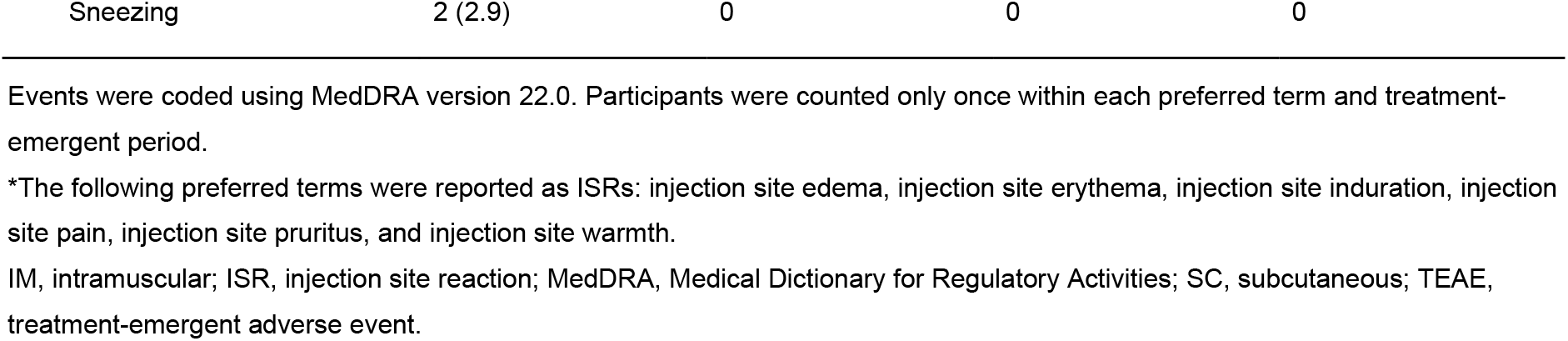
TEAEs and TEAEs Related to Study Treatment (Safety Population).

|  | **Black** |  | **White** |  |
| --- | --- | --- | --- | --- |
|  | Peginterferon beta-1a<br>125 µg SC<br>(*n* = 69) | Peginterferon beta-1a<br>125 µg IM<br>(*n* = 66) | Peginterferon beta-1a<br>125 µg SC<br>(*n* = 58) | Peginterferon beta-1a<br>125 µg IM<br>(*n* = 59) |
| **TEAE, no. (%)** |  |  |  |  |
| Any TEAE | 44 (63.8) | 36 (54.5) | 44 (75.9) | 47 (79.7) |
| Any TEAE related to treatment | 42 (60.9) | 33 (50.0) | 43 (74.1) | 45 (76.3) |
| ISR TEAEs related to treatment* | 14 (20.3) | 6 (9.1) | 26 (44.8) | 11 (18.6) |
| TEAEs related to treatment<br>occurring in >2% of participants<br>by MedDRA Preferred Term |  |  |  |  |
| Chills | 13 (18.8) | 18 (27.5) | 22 (37.9) | 27 (45.8) |
| Headache | 25 (36.2) | 18 (27.3) | 25 (43.1) | 26 (44.1) |
| Injection site erythema | 10 (14.5) | 0 | 22 (37.9) | 2 (3.4) |
| Pain | 6 (8.7) | 11 (16.7) | 12 (20.7) | 17 (28.8) |
| Injection site pain | 8 (11.6) | 4 (6.1) | 10 (17.2) | 9 (15.3) |
| Pyrexia | 8 (11.6) | 4 (6.1) | 2 (3.4) | 8 (13.6) |
| Injection site pruritus | 4 (5.8) | 1 (1.5) | 7 (12.1) | 0 |
| Feeling hot | 1 (1.4) | 3 (4.5) | 6 (10.3) | 4 (6.8) |
| Nausea | 3 (4.3) | 2 (3.0) | 4 (6.9) | 6 (10.2) |
| Back pain | 6 (8.7) | 2 (3.0) | 5 (8.6) | 1 (1.7) |
| Dizziness | 1 (1.4) | 3 (4.5) | 2 (3.4) | 4 (6.8) |
| Decreased appetite | 2 (2.9) | 4 (6.1) | 2 (3.4) | 2 (3.4) |
| Arthralgia | 4 (5.8) | 2 (3.0) | 1 (1.7) | 1 (1.7) |
| Injection site warmth | 2 (2.9) | 0 | 3 (5.2) | 0 |
| Myalgia | 3 (4.3) | 1 (1.5) | 2 (3.4) | 3 (5.1) |
| Pain in extremity | 1 (1.4) | 2 (3.0) | 0 | 3 (5.1) |
| Vomiting | 1 (1.4) | 0 | 1 (1.7) | 3 (5.1) |
| Somnolence | 0 | 3 (4.5) | 1 (1.7) | 1 (1.7) |
| Fatigue | 2 (2.9) | 0 | 2 (3.4) | 0 |
| Flushing | 0 | 0 | 0 | 2 (3.4) |
| Injection site bruising | 0 | 0 | 2 (3.4) | 0 |
| Influenza-like illness | 2 (2.9) | 1 (1.5) | 1 (1.7) | 2 (3.4) |
| Injection site rash | 0 | 0 | 2 (3.4) | 0 |
| Nasal congestion | 1 (1.4) | 1 (1.5) | 2 (3.4) | 1 (1.7) |
| Erythema | 2 (2.9) | 0 | 0 | 0 |
| Eye irritation | 2 (2.9) | 1 (1.5) | 1 (1.7) | 1 (1.7) |
| Injection site induration | 2 (2.9) | 1 (1.5) | 1 (1.7) | 0 |
| Sneezing | 2 (2.9) | 0 | 0 | 0 |

Events were coded using MedDRA version 22.0. Participants were counted only once within each preferred term and treatment-emergent period.
\*The following preferred terms were reported as ISRs: injection site edema, injection site erythema, injection site induration, injection site pain, injection site pruritus, and injection site warmth.
IM, intramuscular; ISR, injection site reaction; MedDRA, Medical Dictionary for Regulatory Activities; SC, subcutaneous; TEAE, treatment-emergent adverse event.

The incidence of TEAEs considered by the study investigator to be related to treatment were similar, with fewer Black than White participants reporting such events with SC (60.9% [42 of 69] vs 74.1% [43 of 58], respectively) or IM (50.0% [33 of 66] vs 76.3% [45 of 59], respectively) administration (Table 2). Incidence of influenza-like illness was similarly low for SC versus IM administration among both Black (2.9% [2 of 69] vs 1.5% [1 of 66]) and White (1.7% [1 of 58] vs 3.4% [2 of 59]) participants. The incidence of injection site reactions (ISRs) related to treatment was greater for SC than IM administration for both Black (20.3% [14 of 69] vs 9.1% [6 of 66], respectively) and White patients (44.8% [26 of 58] vs 18.6% [11 of 59], respectively).

## DISCUSSION

Historically, clinical trials of novel treatments for MS have low numbers of participants who identify as Black, limiting the relevance of available safety and efficacy data to these patients. This is especially concerning regarding MS in the United States, where the Black patient population experiences a higher disease burden than the White patient population.^7^ The recent phase 1 study of IM peginterferon beta-1a demonstrating bioequivalence of the drug administered via SC and IM injection enrolled a similar proportion of Black and White participants,^29^ enabling a subgroup analysis comparing these groups.

Although Black participants displayed slightly higher peginterferon C_max_ and AUC values compared with White participants, other PK and PD values were similar between groups. Additionally, no differences between groups were observed for either PD or safety outcomes. The incidence of TEAEs and TEAEs related to treatment following IM or SC administration in Black participants did not exceed, and appeared to be lower than, the incidence in White participants. Most common reported TEAEs related to treatment were similar in both groups, with chills and headache reported most frequently. No SAEs were observed in either group.

Thus, PK, PD, and safety outcomes following peginterferon beta-1a SC or IM administration were generally similar between Black and White study participants. The differences in C_max_ and AUC between groups did not result in greater safety concerns or differences in neopterin levels or dynamics, suggesting no clinically meaningful differences and thus no need for changes in dosage based on race.

To our knowledge, this study is the first to directly compare PK and PD profiles of an interferon or other MS DMT between racial groups. Previous clinical studies of beta interferons have suggested that Black patients with MS may have a less favorable treatment response to beta interferons compared with White patients. A post hoc analysis of the EVIDENCE clinical trial found that Black patients had more relapses and magnetic resonance imaging (MRI) disease activity after 24 and 48 weeks of treatment with interferon beta-1a compared with White patients; however, the number of non-White participants was limited.^9^ A single-center, cross-sectional study of MRI characteristics in a sample of Black and White patients with similar MS disease durations, most of whom were treated with beta interferons (71% of Black and 68% of White) reported that Black patients had significantly greater T1 and T2 lesion volumes than White patients, suggesting poorer disease activity control in the former group.^11^ Evidence is mixed from studies of other DMTs, including high-efficacy DMTs such as natalizumab and ocrelizumab, with some studies suggesting a poorer response to treatment in Black compared with White patients,^26^ and others reporting a finding of no difference.^10, 12, 31^ Interpretation of the previous studies is limited by the low proportion of Black patients included, which ranged from 1.9% to 13.9%. In contrast, this study enrolled greater than 50% Black participants, alongside a similar proportion of White participants. Such comparable proportions in the field of MS have historically been limited to retrospective chart reviews.^26^ Further research is needed to better understand observed differences across patient populations with MS. Such studies may also shed light on differences in therapeutic effect in patients with MS.

Inequity in healthcare, including underrepresentation of certain racial and ethnic groups in clinical studies, is recognized as a significant problem.^1, 3, 32, 33^ Underrepresented racial and ethnic groups face specific barriers to participation and retention in clinical studies including, but not limited to, a lack of information on relevant clinical studies, biases on the part of referring physicians or study investigators, and a lack of trust in the medical establishment.^4, 13, 34, 35^ The consequences of lack of representation in clinical studies include limited generalizability of the results and inaccuracies in racial-or ethnicity-specific subgroup analyses.^13^ These can, in turn, lead to a poor translation from the clinic into real-world use and over-or underestimation of the risks or benefits of medications.

In an analysis of 167 drugs approved by the US Food and Drug Administration from 2008–2013, approximately 20% demonstrated racial and/or ethnic differences in exposure and/or therapeutic response.^36^ Moving forward, it will be essential to include socioeconomic determinants and other potentially conflating factors to determine which of these treatments produce differences in response that are independently associated with race. Continued development of guidelines and best practices for the recruitment and retention of underrepresented populations in clinical studies of MS DMTs^15, 37^ is of critical importance to understand and address the persistent problems in health equity faced by these patients.

A thorough understanding of the impact of race and ethnicity on the biological properties of MS DMTs in Black patients as well as patients from other underrepresented racial or ethnic groups is needed and will be aided by studies specially designed to assess PK, PD, safety, and treatment effects in all relevant patient populations. Inclusion of sufficient numbers of relevant populations in clinical studies is essential to accurately evaluate safety and response to therapy. These efforts may be aided by selecting clinical trial recruitment sites in areas with balanced demographics, and including investigators who reflect the diversity of the patient population.^17^

The findings from this post hoc analysis suggest there is no meaningful clinical difference between Black and White participants in the PK, PD, and safety response to peginterferon beta-1a administered by SC or IM injection. Additional studies of peginterferon beta-1a in well-balanced racial and ethnic cohorts of patients with MS will be needed to confirm this conclusion in the therapeutic setting.

## Data Availability

The datasets generated and/or analyzed during the current study are not publicly available. The authors fully support sharing whenever possible. Requests for de-identified data should be made to Biogen via established company data-sharing policies and processes detailed on the website http://clinicalresearch.biogen.com/.

## ACKNOWLEDGMENTS

Medical writing and editorial support for the development of this manuscript, under the direction of the authors, were provided by Ashfield MedComms, an Ashfield Health company. Funding for medical writing support was provided by Biogen Inc. (Cambridge, MA, USA).

## CONTRIBUTIONS

C.L.B., Y.Z., M.L.N., and N.R. designed the bioequivalence study and this analysis. Y.Z. and O.M. were responsible for the statistical analysis. All authors had access to the data, contributed to data interpretation, and attest to its completeness and accuracy. All authors contributed to the writing and review of the manuscript, and assume responsibility for the integrity of the work as a whole, including the accuracy of the data. All authors have read and approved the final draft,, and agreed to submit the manuscript for publication.

## SUPPLEMENTARY APPENDIX

**Table S1.**
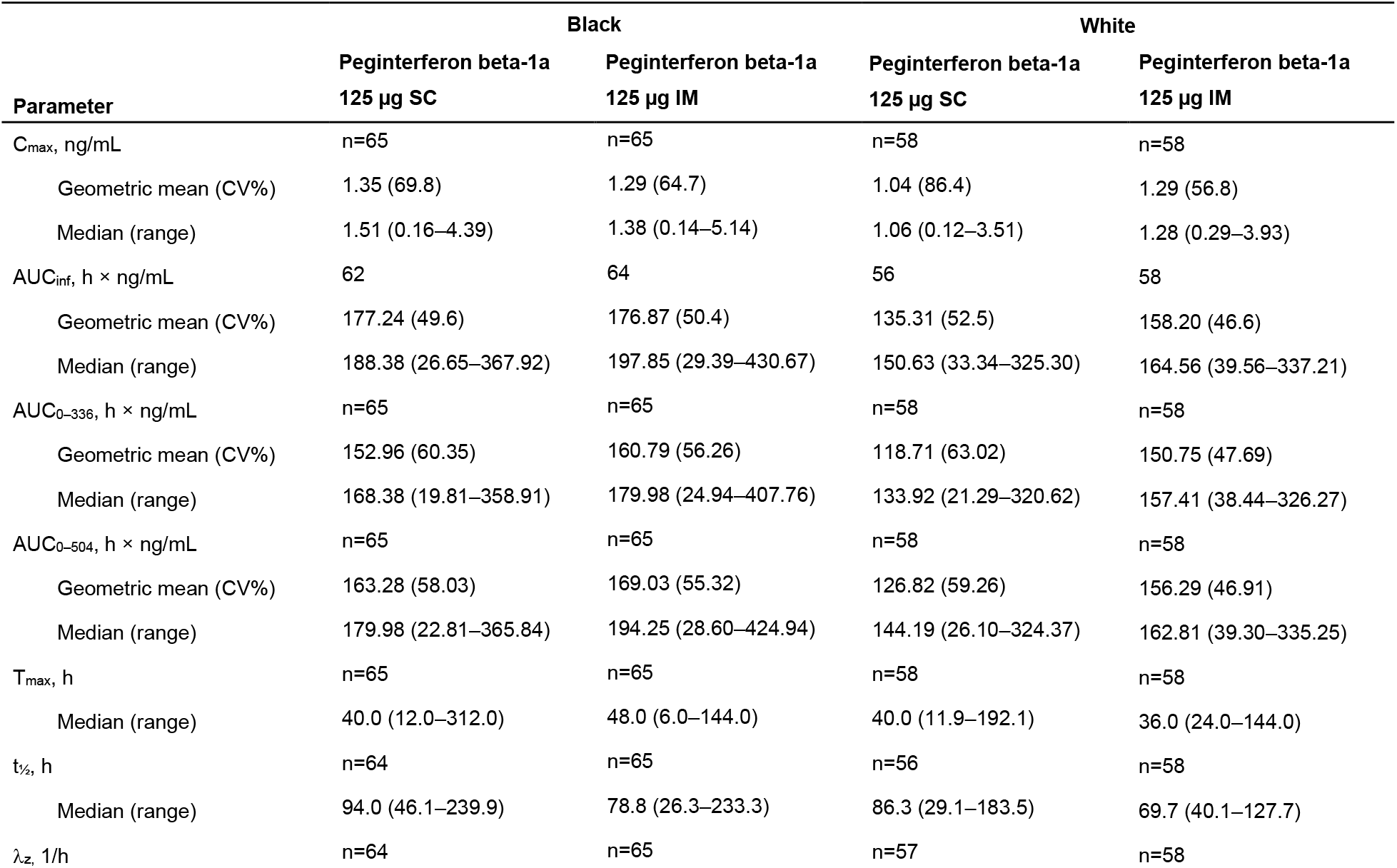
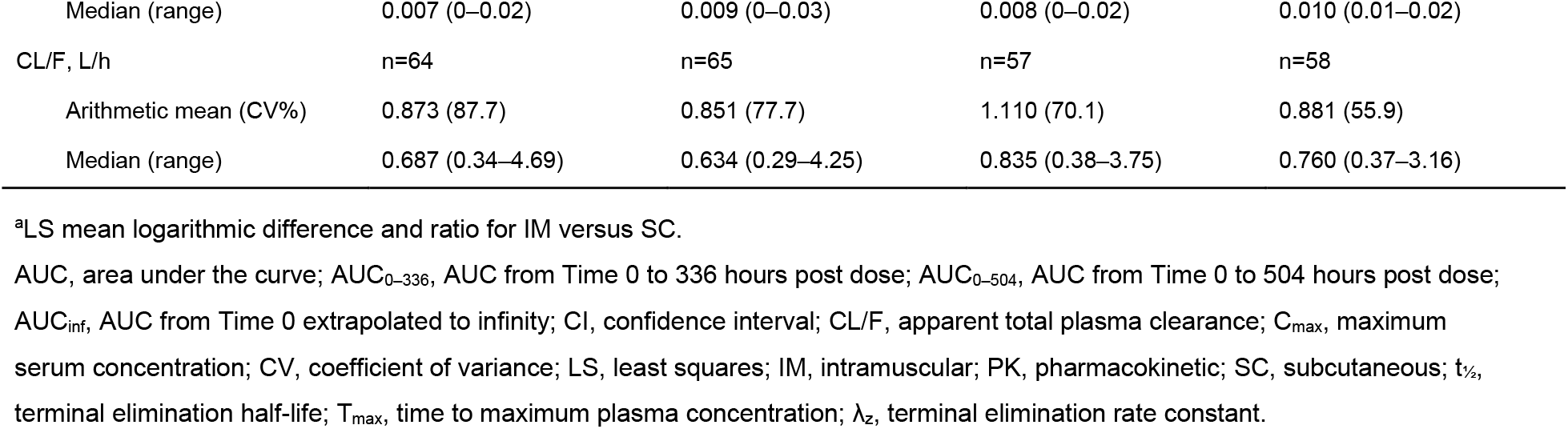
Pharmacokinetic parameters of peginterferon beta-1a following IM and SC administration (PK population).

| Parameter | Black |  | White |  |
| --- | --- | --- | --- | --- |
|  | Peginterferon beta-1a<br>125 µg SC | Peginterferon beta-1a<br>125 µg IM | Peginterferon beta-1a<br>125 µg SC | Peginterferon beta-1a<br>125 µg IM |
| C<sub>max</sub>, ng/mL | n=65 | n=65 | n=58 | n=58 |
| Geometric mean (CV%) | 1.35 (69.8) | 1.29 (64.7) | 1.04 (86.4) | 1.29 (56.8) |
| Median (range) | 1.51 (0.16–4.39) | 1.38 (0.14–5.14) | 1.06 (0.12–3.51) | 1.28 (0.29–3.93) |
| AUC<sub>inf</sub>, h × ng/mL | 62 | 64 | 56 | 58 |
| Geometric mean (CV%) | 177.24 (49.6) | 176.87 (50.4) | 135.31 (52.5) | 158.20 (46.6) |
| Median (range) | 188.38 (26.65–367.92) | 197.85 (29.39–430.67) | 150.63 (33.34–325.30) | 164.56 (39.56–337.21) |
| AUC<sub>0–336</sub>, h × ng/mL | n=65 | n=65 | n=58 | n=58 |
| Geometric mean (CV%) | 152.96 (60.35) | 160.79 (56.26) | 118.71 (63.02) | 150.75 (47.69) |
| Median (range) | 168.38 (19.81–358.91) | 179.98 (24.94–407.76) | 133.92 (21.29–320.62) | 157.41 (38.44–326.27) |
| AUC<sub>0–504</sub>, h × ng/mL | n=65 | n=65 | n=58 | n=58 |
| Geometric mean (CV%) | 163.28 (58.03) | 169.03 (55.32) | 126.82 (59.26) | 156.29 (46.91) |
| Median (range) | 179.98 (22.81–365.84) | 194.25 (28.60–424.94) | 144.19 (26.10–324.37) | 162.81 (39.30–335.25) |
| T<sub>max</sub>, h | n=65 | n=65 | n=58 | n=58 |
| Median (range) | 40.0 (12.0–312.0) | 48.0 (6.0–144.0) | 40.0 (11.9–192.1) | 36.0 (24.0–144.0) |
| t<sub>1/2</sub>, h | n=64 | n=65 | n=56 | n=58 |
| Median (range) | 94.0 (46.1–239.9) | 78.8 (26.3–233.3) | 86.3 (29.1–183.5) | 69.7 (40.1–127.7) |
| λ<sub>z</sub>, 1/h | n=64 | n=65 | n=57 | n=58 |
| Peginterferon beta-1a PK/PD by Race – Supplemental Information |  |  |  |  |
| Median (range) | 0.007 (0–0.02) | 0.009 (0–0.03) | 0.008 (0–0.02) | 0.010 (0.01–0.02) |
| CL/F, L/h | n=64 | n=65 | n=57 | n=58 |
| Arithmetic mean (CV%) | 0.873 (87.7) | 0.851 (77.7) | 1.110 (70.1) | 0.881 (55.9) |
| Median (range) | 0.687 (0.34–4.69) | 0.634 (0.29–4.25) | 0.835 (0.38–3.75) | 0.760 (0.37–3.16) |

<sup>a</sup>LS mean logarithmic difference and ratio for IM versus SC.
AUC, area under the curve; AUC<sub>0–336</sub>, AUC from Time 0 to 336 hours post dose; AUC<sub>0–504</sub>, AUC from Time 0 to 504 hours post dose; AUC<sub>inf</sub>, AUC from Time 0 extrapolated to infinity; CI, confidence interval; CL/F, apparent total plasma clearance; C<sub>max</sub>, maximum serum concentration; CV, coefficient of variance; LS, least squares; IM, intramuscular; PK, pharmacokinetic; SC, subcutaneous; t<sub>1/2</sub>, terminal elimination half-life; T<sub>max</sub>, time to maximum plasma concentration; λ<sub>z</sub>, terminal elimination rate constant.

**Table S2.**
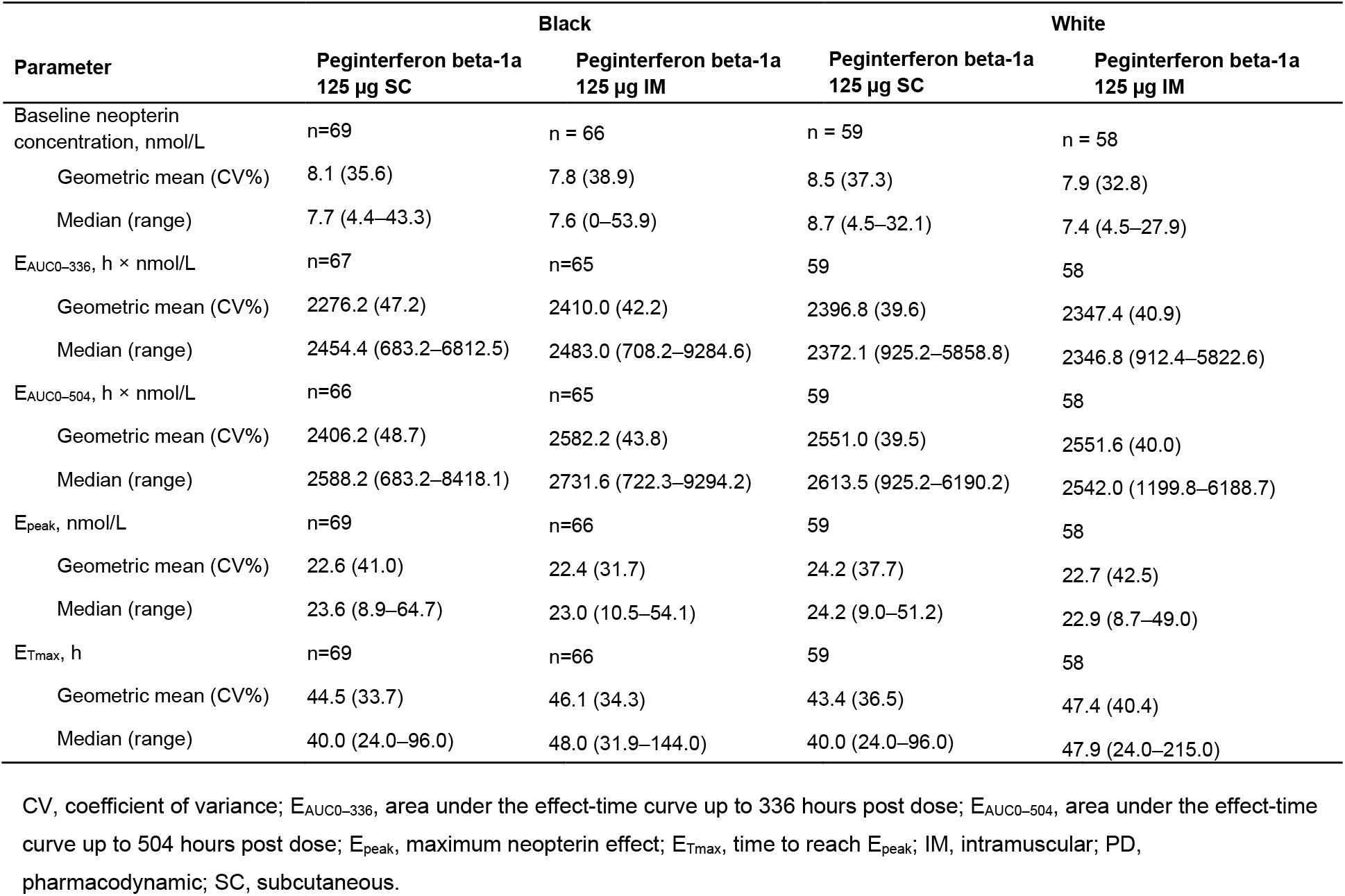
Pharmacodynamic parameters of peginterferon beta-1a following IM and SC administration (PD population).

